# SURPASS-HF: Safety and Utility of Remote Pulmonary Artery Sensor Shared-management in Heart Failure

**DOI:** 10.64898/2026.07.10.26357468

**Authors:** Marc Atzenhoefer, Hussain Boxwala, Tamar Atzenhoefer, Michelle Staudacher, Fahad Iqbal

**Author notes:** Corresponding author: Marc Atzenhoefer, MD —. Ethics & oversight: Approved by the Froedtert/MCW IRB (protocol #PRO00056040) before the 14 January 2026 start, per the Declaration of Helsinki. All participants gave written informed consent that disclosed the risks of patient-executed diuretic up-titration (volume depletion, AKI, electrolyte disturbance, hypotension/falls). Funding: None.

## Abstract

**Background:** Insulin-dependent diabetics self-titrate therapy to self-obtained glucose values as standard of care, yet heart failure (HF) patients with implanted pulmonary artery (PA) pressure sensors never see their own readings; clinicians interpret and execute every dose change - a model that does not scale to a ∼200-patient HF panel. To our knowledge, SURPASS-HF is the first prospective feasibility study applying the insulin-titration paradigm to PA-pressure-guided HF care: patients executing a prescribed loop-diuretic sliding scale, supported by ARTHUR, a domain-trained large language model, with clinician confirmation of every adjustment.

**Methods:** Non-randomized, prospective, single-arm, single-center 90-day feasibility study (January 14-April 14, 2026; 60.1 patient-months). Twenty-one adults with implanted PA sensors enrolled (intention-to-treat, ITT); 19 completed full follow-up (per-protocol, PP). Regimens and individual PA diastolic (PAD) targets were explicitly prescribed; when daily pressures met published serial-reading thresholds, the software prepared the pre-determined adjustment, the clinician confirmed it, and the patient executed it. ARTHUR reinforced dose ceilings, prompted surveillance labs, and escalated edge cases for review. Pre-specified outcomes: adverse events, escalations, time in optimal PA range (TIR-PAP, +/-5 mmHg of goal), reading adherence, provider overrides, and paired delta_PAD (first vs last 7-day windows). Confidence intervals are descriptive; the study was not powered for significance.

**Results:** Mean age was 69+/-11 years, 52% women, mean baseline PAD 14.8 mmHg. No pre-specified safety event (KDIGO >or=1 AKI, hyperkalemia, hyponatremia, symptomatic hypotension) was detected (0/8 post-adjustment draws in 5/21 patients; exact 95% CI 0-37%); laboratory ascertainment was sparse, so a meaningful harm rate cannot be excluded. Seventeen of 19 PP patients (89%) required no protocol-triggered escalation; 4 escalations occurred in 2 patients. TIR-PAP was 88.4% (ITT)/91.3% (PP); reading adherence 92.1%; 53 provider alerts (0.88/patient-month) all resolved (median 24 h) with no overrides. delta_PAD was -0.89 mmHg (ITT; 95% CI -2.60 to +0.82) in a cohort already at goal at baseline. Two non-cardiac hospitalizations occurred.

**Conclusions:** LLM-mediated, clinician-confirmed patient execution of a published deterministic PA-pressure-guided diuretic algorithm was feasible over 90 days, with high time-in-range and adherence and no detected safety events. Findings from this prospective, single arm, non-randomized, small cohort are descriptive. The study was not designed or powered to demonstrate evidence of a treatment effect; a randomized, well powered prospective comparison study against provider-led PA-pressure management is the next ideal step.

## Introduction

A patient with type I diabetes adjusts their dose of insulin multiple times per day to measurements they obtain themselves. Primary care providers (PCP) regard this strategy as standard of care. Compare this to the heart failure (HF) patient with an implanted pulmonary artery (PA) pressure sensor. Both patients produce equally precise and actionable metrics, however the HF patient never sees their value. Upwards of a week, they measure daily pressures and wait for their clinician to read, interpret, and communicate medication changes, if necessary, through a complex care delivery apparatus. The asymmetry extends beyond its structure and execution. The cardiologist’s HF panel (∼200 patients) dwarfs that of the average PCP’s insulin-dependent diabetic population (∼16). The prevailing provider-dependent treatment strategy in the face of 6.7 million Americans with HF^1^ is simply unrealistic (Figure 1). A published field guide^2^ specifies the targets and threshold rules for PA-pressure–guided diuretic management, which we have operationalized in our community HF program (SMART-HF; preprint^3^). Structured management reduces HF hospitalizations^4^ reliably achieving pressure deltas that track with reductions in mortality. ^5^ Unfortunately, precision HF management reaches a fraction of eligible patients. In the trials founding this strategy (CHAMPION^4^), clinicians executed every adjustment. To our knowledge, SURPASS-HF is the first prospective test of the insulin therapy paradigm to PA-pressure–guided HF: patients executing a prescribed loop-diuretic sliding scale. Facilitating the otherwise complex care delivery layer, ARTHUR, a domain-trained large language model supported patient communication, execution, and safety monitoring strictly according to our usual structured algorithmic approach.

**Figure 1:**
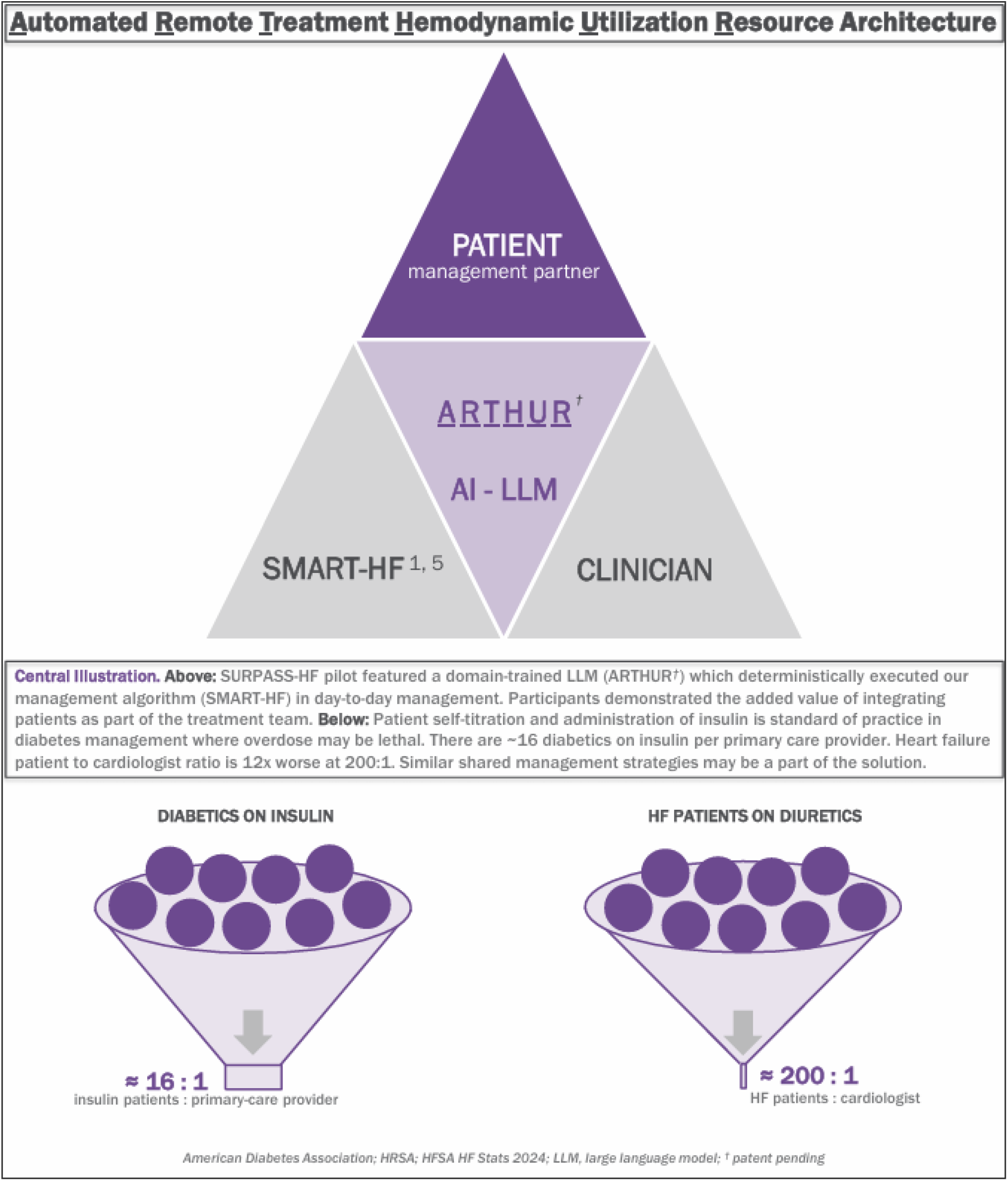
Central Illustration. *Above:* SURPASS-HF model visualized. Patients receive PA pressures with an analysis from ARTHUR (a domain-trained LLM) daily with, if indicated, detailed/individualized change(s) to treatment plan according to the deterministic clinician pre-programmed, threshold-gated decision logic. *Below:* The clinician-burden gap compared. ∼16 insulin-treated patients per primary-care provider versus ∼200 HF patients per cardiologist. Burden figures are illustrative estimates from HFSA HF STATS 2024, the American Diabetes Association, and HRSA.

## Methods

Non-randomized, prospective, single-arm, single-center feasibility study (14 January–14 April 2026; 90 days; 60.1 patient-months). Twenty-one patients with implanted PA pressure sensors enrolled (intention-to-treat, ITT); 19 completed full follow-up (per-protocol, PP); two late enrollers (days 31, 41) were ITT-only. Each patient’s medication regimen and individual PAD target^2^ were explicitly prescribed. This regimen changed only when daily pressures met published serial-reading thresholds^2^, whereupon the software prepared the appropriate pre-determined adjustment, the clinician confirmed it, and the patient subsequently executed it. ARTHUR reinforced dose ceilings prompted post-dose adjustment lab draws, and escalated edge cases to the provider’s attention when necessary. Pre-specified outcomes: adverse events; escalation count; time in optimal PA range (TIR-PAP, ±5 mmHg of goal); reading adherence; manual provider overrides; and paired ΔPAD (first vs last 7-day windows during 90-day observation period) Study was not powered for significance; CIs are descriptive.

## Results

Three pre-defined groups enrolled: 16 well-controlled (PAD target <20 mmHg; ≤1 adjustment/3 months pre-study), 3 poorly controlled (target ≥20 mmHg; historically frequent adjustments required), and 2 new implants. The cohort began at goal (mean baseline PAD 14.8 mmHg), mean age of 69±11 years and 52% women (11/21). No pre-specified safety events detected: zero KDIGO stage ≥1 AKI, hyperkalemia, hyponatremia, or symptomatic hypotension. Note, post escalation laboratory studies were sparse (8 draws in 5/21 patients statistically limited and thus a meaningful harm rate cannot be excluded.

Seventeen of 19 PP patients (89%) required zero protocol triggered diuretic escalation; 4 escalations triggered by 2 patients (PAD ≥6 mmHg above target on three serial readings^2^). One of these escalations precipitated by prednisone prescribed for COPD exacerbation in a patient with chronic hypoxemic respiratory failure on nasal cannula oxygen. The remaining three events were attributable to behavioral/dietary indiscretion. TIR-PAP was 88.4% (ITT)/91.3% (PP); reading adherence 92.1%; provider alerts 0.88/patient-month (53; all resolved, median 24 h; no overrides). The pre-specified ΔPAD was −0.89 mmHg (ITT; 95% CI −2.60 to +0.82): the cohort mean did not rise, descriptive but not powered for equivalence and furthermore in an already-at-goal cohort. A post-hoc above-median-baseline subgroup (n=11/21) shifted −2.34 mmHg. Two non-cardiac hospitalizations occurred (disequilibrium [unrelated] and COPD exacerbation).

## Discussion

SURPASS-HF shows that LLM-mediated, clinician-confirmed delivery of a published deterministic PA-pressure algorithm^2^ is feasible: over 60.1 patient-months, 89% of PP patients needed no escalation, TIR was 88–91%, and no adverse events were detected. With no shortage of limitations, these findings outline feasibility of execution for well-controlled HF patients and without evidence of a treatment effect. Superiority and safety at scale await well powered and randomized comparison studies. A domain-trained LLM can render an established deterministic HF algorithm executable as a patient shared-management strategy potentially extending precision hemodynamic-guided HF care beyond the limited capacity of current provider-only execution methodology (CHAMPION^4^). We extended a widely accepted paradigm leveraged in diabetes management to a select HF population. Limitations: small, single-center, single-arm; nearly entire cohort already at goal; only 90-day follow-up data thus far. A randomized, powered trial against ad-hoc provider-led PA-pressure management with hard endpoints like HF hospitalizations, PA-pressure deltas, and extended follow up is the next step.

## Data Availability

All de-identified relevant data in this study are available upon request from the authors

## Notes

Regulatory: ARTHUR (domain trained LLM) functioned solely as clinician support and execution tool. Every therapeutic adjustment was pre-determined and specified. Zero autonomous treatment decisions executed.

Disclosures: M.A. authored the algorithm deployed by ARTHUR. under study (refs 2,3) and is a consultant/speaker for Abbott Cardiovascular (CardioMEMS regional KOL); Abbott had no role in design, analysis, or the decision to submit. ARTHUR is patent pending. Remaining authors report no conflicts.

### Competing Interest Statement

Marc Atzenhoefer, MD is a KOL for the Abbott CardioMEMS HF system.

### Author Declarations

Ethics: Medical College of Wisconsin Informed Consent for Research (MCW IRB) gave ethical approval for this work (Minimal risk IRB protocol # PRO00056040), Declaration of Helsinki, written informed consent.

